# An efficient distributed algorithm with application to COVID-19 data from heterogeneous clinical sites

**DOI:** 10.1101/2020.11.17.20220681

**Authors:** Jiayi Tong, Chongliang Luo, Md Nazmul Islam, Natalie Sheils, John Buresh, Mackenzie Edmondson, Peter A. Merkel, Ebbing Lautenbach, Rui Duan, Yong Chen

## Abstract

**Objectives:** Integrating electronic health records (EHR) data from several clinical sites offers great opportunities to improve estimation with a more general population compared to analyses based on a single clinical site. However, sharing patient-level data across sites is practically challenging due to concerns about maintaining patient privacy. The objective of this study is to develop a novel distributed algorithm to integrate heterogeneous EHR data from multiple clinical sites without sharing patient-level data.

**Materials and Methods:** The proposed distributed algorithm for binary regression can effectively account for between-site heterogeneity and is communication-efficient. Our method is built on a pairwise likelihood function in the extended Mantel-Haenszel regression, which is known to be statistically highly efficient. We construct a surrogate pairwise likelihood function through approximating the target pairwise likelihood by its surrogate. We show that the proposed surrogate pairwise likelihood leads to a consistent and asymptotically normal estimator by effective communication without sharing individual patient-level data. We study the empirical performance of the proposed method through a systematic simulation study and an application with data of 14,215 COVID-19 patients from 230 clinical sites at UnitedHealth Group Clinical Research Database.

**Results:** The proposed method was shown to perform close to the gold standard approach under extensive simulation settings. When the event rate is <5%, the relative bias of the proposed estimator is 30% smaller than that of the meta-analysis estimator. The proposed method retained high accuracy across different sample sizes and event rates compared with meta-analysis. In the data evaluation, the proposed estimate has a relative bias <9% when the event rate is <1%, whereas the meta-analysis estimate has a relative bias at least 10% higher than that of the proposed method.

**Conclusions:** Our simulation study and data application demonstrate that the proposed distributed algorithm provides an estimator that is robust to heterogeneity in event rates when effectively integrating data from multiple clinical sites. Our algorithm is therefore an effective alternative to both meta-analysis and existing distributed algorithms for modeling heterogeneous multi-site binary outcomes.

## INTRODUCTION

Electronic health records (EHR) data have become one of the most well-known data sources for medical and health research use. EHRs contain various elements of patient-level health information, including diagnoses, medications, procedures, imaging, and clinical notes [1-4]. Synthesis of this real-world evidence (RWE) from multiple clinical sites provides a larger sample size of the population compared to a single site study [5]. Analyses using larger populations benefit from better accuracy in estimation and prediction. Furthermore, the integration of research networks inside healthcare systems allows rapid translation and dissemination of research findings into evidence-based healthcare decision making to improve health outcomes, consistent with the idea of a learning health system [6-11].

In the past few years, several successful networks have been founded and become beneficial to multicenter research. One of them is the Observational Health Data Sciences and Informatics (OHDSI) consortium [12]. OHDSI was founded for the primary purpose of developing open-source tools that could be shared across multiple sites. OHDSI developed the Observational Medical Outcomes Partnership (OMOP) Common Data Model (CDM) for data standardization [13]. The OMOP allows each institution to transform the local EHR data to the CDM’s standards. This procedure makes it feasible for the researchers to develop methods that can be simultaneously applied to the datasets from many institutions. The conversion and standardization of the data format decrease the probability of translation error and also increase the efficiency of data analysis. Another successful network is the National Pediatric Learning Health System (PEDSnet), a National Pediatric Learning Health System, within the PCORnet system [14,15]. This network contains eight large pediatric health systems in the US. Comprising clinical information from millions of children, PEDSnet offers the capacity to conduct multicenter pediatric research with broad real-world evidence. The Sentinel System is another example of a multi-site network, a national electronic system for monitoring performance of FDA-regulated medical products [16].

In multi-center studies, maintaining privacy of patient data is a major challenge [17-19]. Due to data privacy policies, directly sharing patient-level data, especially demographic, comorbidity, and outcome data, is restricted and poorly feasible in practice. The Health Insurance Portability and Accountability Act of 1996 (HIPAA) introduced a privacy rule to regulate use of protected health information (PHI) often found in EHRs, requiring de-identification of PHI before use in biomedical research [18]. De-identified PHI has been shown to be susceptible to re-identification, causing concern among patients [20,21].

In light of patient privacy concerns, many multicenter EHR-based studies currently conduct analyses by combining shareable summary statistics through meta-analysis [22-24]. While relatively simple to use, meta-analysis has been shown to result in biased or imprecise estimation in the context of rare outcomes, as well as with smaller sample sizes [25]. Other than meta-analysis, several distributed algorithms have been developed and considered in studies with multi-site data. In these distributed algorithms, a model estimation process is decomposed into smaller computational tasks that are distributed to each site. After parallel computation, intermediate results are transferred back to the coordinating center for final synthesis. Under this framework, there is no need to share patient-level data across sites. For example, GLORE (Grid Binary LOgistic Regression) was developed for conducting distributed logistic regressions [26], and WebDISCO (a Web service for distributed Cox model learning) was developed to fit the Cox proportional hazard model distributively and iteratively [27]. Both algorithms have been successfully deployed to the pSCANNER consortium [28]. Through iterative communication of aggregated information across the sites, these two algorithms provide accurate and lossless results, which are equivalent to fitting a model on the pooled data from all sites. However, in practice these two methods can be time-consuming and communication-intensive due to the need for iteratively transferring data. To overcome this limitation, non-iterative privacy-preserving distributed algorithms for logistic regression (termed as ODAL) and Cox model (termed as ODAC) through the construction of a surrogate likelihood have been proposed [25,29,30].

However, all of the aforementioned distributed algorithms rely on the assumption that data across clinical sites are homogeneous. This assumption is often inaccurate in biomedical studies because it ignores the heterogeneity caused by the intrinsic differences across clinical sites or population characteristics. Ignoring heterogeneity across clinical sites can induce biases in estimating associations between the exposures of interest and outcomes [17,31]. Recently, a single Robust-ODAL algorithm was proposed to account for the heterogeneity across the clinical sites, but it only considers the limited situation when there exist a small number of outlying studies within the network [32].

One motivating example is the EHR data of 14,215 patients who were diagnosed with COVID-19 prior to June 29, 2020 from 230 sites in the UnitedHealth Group Clinical Research Database. There is a substantial difference in clinical practices across these sites due to such factors as geographical variability in disease patterns, variations in patients’ characteristics, and regional differences in practice patterns. Therefore, developing methods to account for the heterogeneity in the data is especially needed when analyzing multi-site data within the networks.

To fill the above methodology gap, in this paper, we develop an effective privacy-preserving distributed data integration algorithm. We propose a distributed algorithm for binary regression to account for between-site heterogeneity by efficient communication (i.e., only requires one round of communication of aggregated information among the sites). Influenced by the pairwise likelihood in the extended Mantel-Haenszel regression [31] and the idea proposed by Jordan et al. (2019) [33], we construct a surrogate pairwise likelihood through approximating the target pairwise likelihood by its surrogate. We show that the proposed surrogate pairwise likelihood leads to a consistent and asymptotically normal estimator, which is asymptotically equivalent to the maximum pairwise likelihood estimator based on the pooled data. This result is established based on U statistics, which is different from Jordan et al. (2019) [33]. We evaluate the empirical performance of the proposed method through simulation studies and apply the proposed method to investigate the associations between length of stay and the risk factors of interests. **Figure 1** shows the comparisons between the pooled analysis, meta-analysis method, iterative distributed algorithms, and the proposed method from various aspects. The proposed method can retain high estimation accuracy, protect patient privacy, handle heterogeneity, and save communication cost compared to the others.

**Figure 1:**
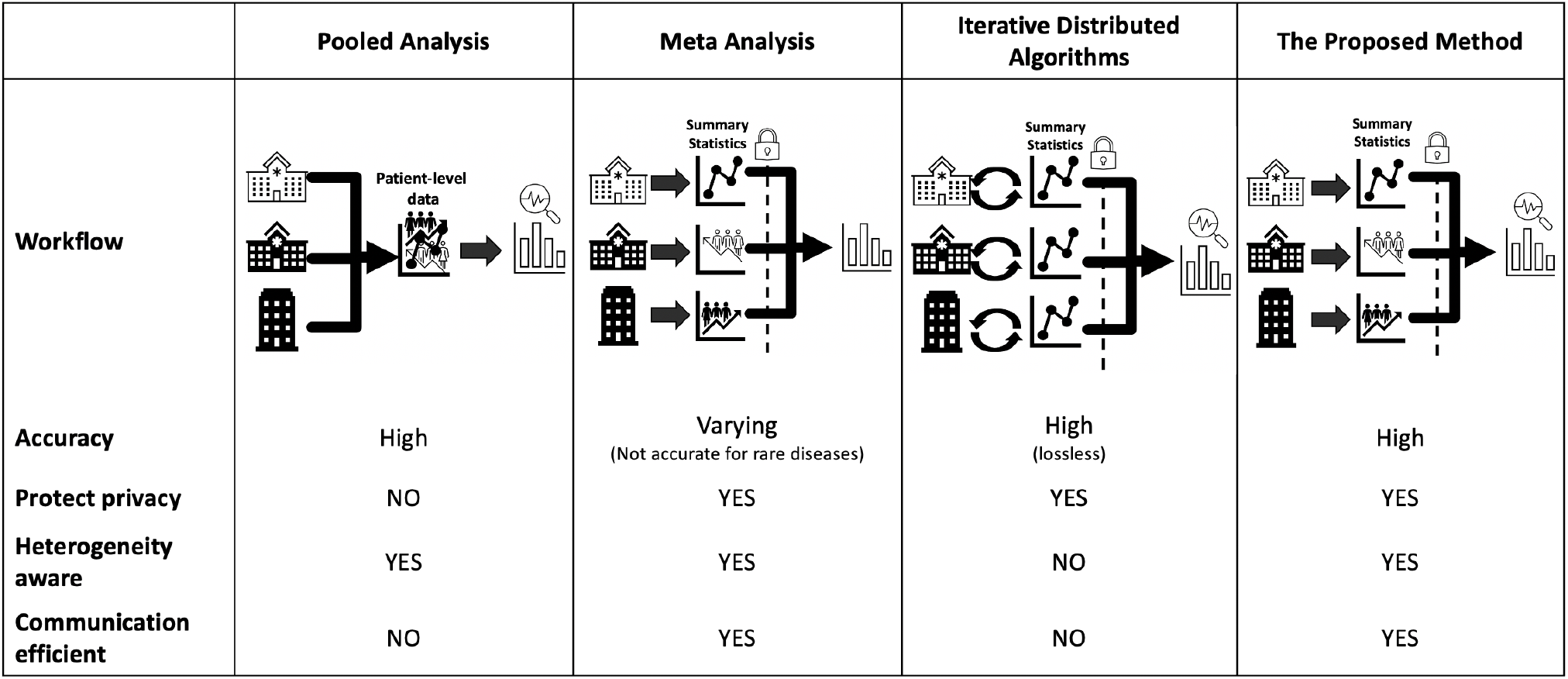
Comparisons between pooled analysis, meta-analysis, iterative distributed algorithms, and the proposed method. The proposed method can retain high accuracy when estimating association between exposures and outcome of interest. In addition, the proposed method can handle heterogeneity across the sites and protect patient privacy with efficient communication.

## MATERIALS AND METHODS

### Surrogate Pairwise Likelihood

Suppose we have K different clinical sites. To keep the notation simple, we assume that each site has an equal number of n patients. Let {*x*_*ij*_} denote the collection of risk factors and {*y*_*ij*_} denote the independent response variable for the j-th patient in the i-th site where *j* = 1,…, *n* and *i*= 1,…, *k*. The logistic regression model to characterize the association between the risk factors and the outcome is

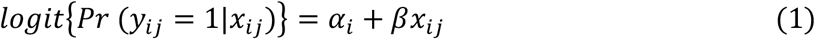

where *logit*(*p*) =*log*{*p*/(1 − *p*)},*α*_*i*_ represents the site-specific prevalence of response variable, and *β* is the log odds ratio, meaning the association between risk factors *x*_*ij*_ and the outcome *y*_*ij*_.

Following Liang (1987)’s extended Mantel-Haenszel regression, the pairwise likelihood can be constructed by conditioning (*y*_*ij;*_ *y*_*il*_) on their order statistics. The pairwise likelihood for the i-th site can be written as

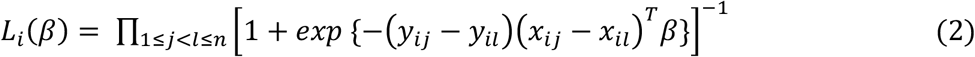

We note that unlike generalized linear mixed effect model, where site-specific effects *α*_*i*_’s are assumed to follow a known distribution, the conditional pairwise likelihood eliminates the nuisance parameters (*α*_*1*_,…, *α*_*k*_) through the conditioning technique, hence avoids estimation of the nuisance parameters. Moreover, as studied in Liang (1987), the estimator, defined as the maximum of the pairwise likelihood, retains high statistical efficiency.

Now summing over all K sites, the overall likelihood function can be written as the product of *L*_*i*_,

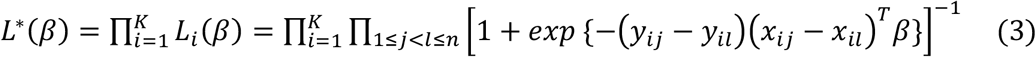

which can be calculated if we have access to the patient-level data from all sites.

However, in practice, the individual patient-level data are only available at the local site and for the rest of the clinical sites in the network, we can only access aggregated information. Motivated by the surrogate likelihood in Jordan et al. (2019) which approximates the target pairwise likelihood by the likelihood from a single site, we propose a surrogate pairwise likelihood which can still handle the heterogeneity across the clinical sites.

For simplicity, we assume the first site as the local site, where we have access to the individual patient-level data. Let *l*_1_(*β*) denote the pairwise log-likelihood function for the local site,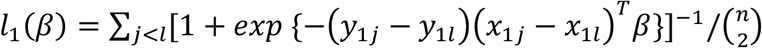. We construct the following surrogate log pairwise likelihood function 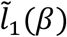 with the patient-level data from the local site, the initial value 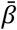, and the aggregated information 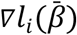 and 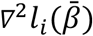. Specifically, we define

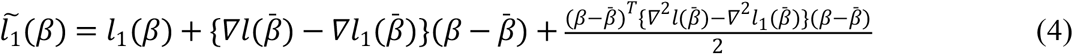

where *l*_1_ (*β*) is the log pairwise likelihood function calculated from patient-level data in the local site; 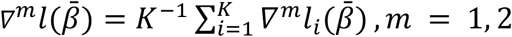 and 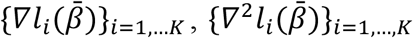 are the first and second gradients of the surrogate pairwise likelihood function at 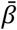 respectively. By maximizing the surrogate pairwise likelihood 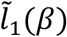 we obtain the surrogate estimator 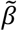.

A natural choice of the initial value of 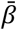 is the maximum likelihood estimator of the local site *l*_1_ (*β*). Alternatively, since the performance of the surrogate estimator 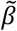 may depend on the choice of the initial values, we may use the inverse variance weighted average of the estimates from all sites, i.e.,

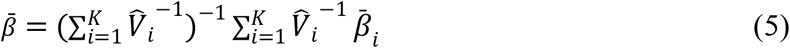

where 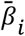 is the maximum of the pairwise likelihood and 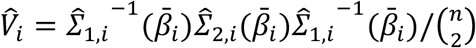 is the covariance matrix of 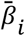 in the i-th site; 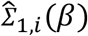 and 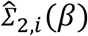 are functions of *β* for the i-th site. The definition of the covariance matrix, the asymptotic distribution of the surrogate estimator 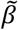, the derivation of the limiting distribution of 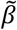 are provided in Supplementary Appendix 1.

#### Algorithm

The proposed surrogate pairwise likelihood leads to the following algorithm.

**Algorithm: Proposed method**

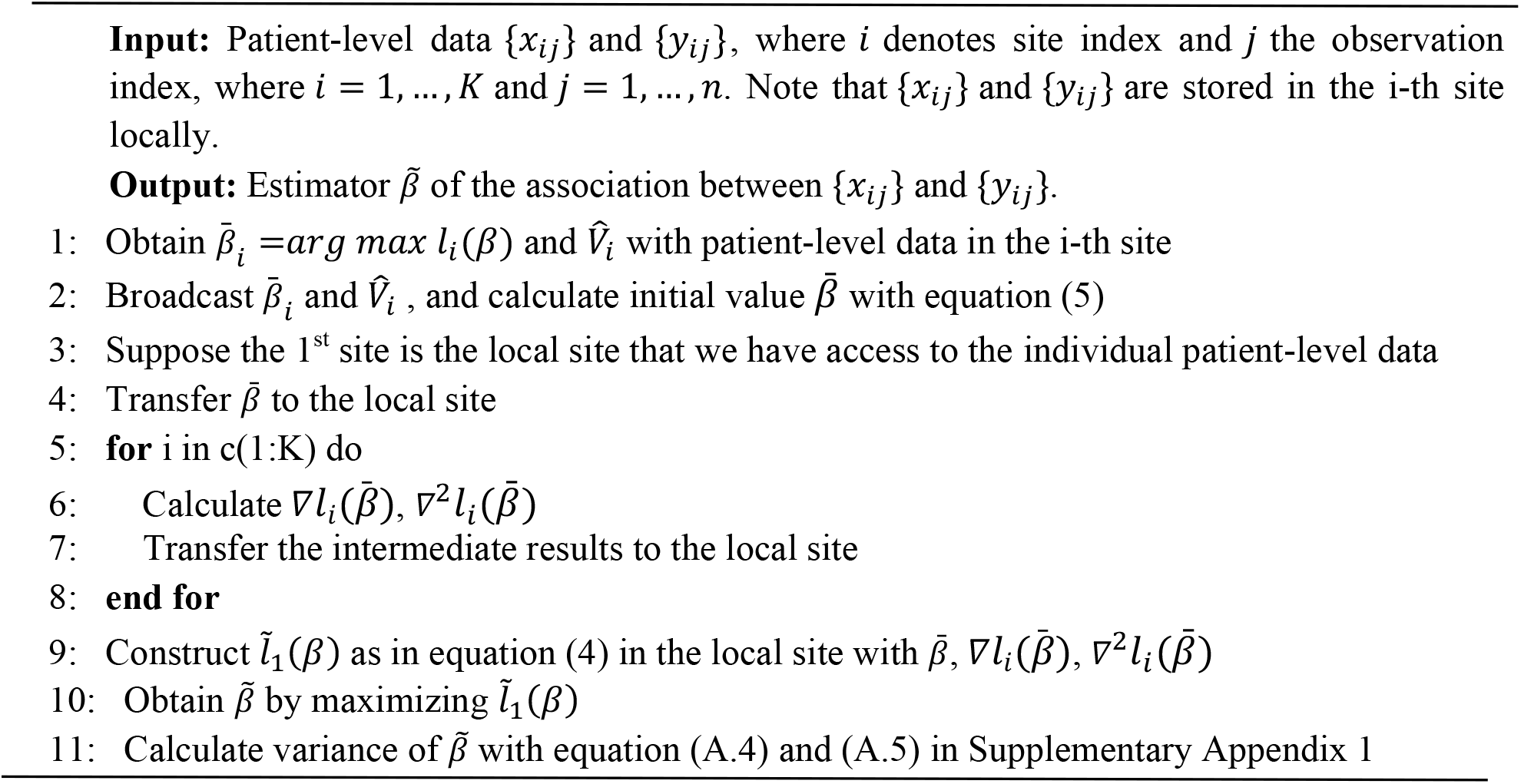

In the following figure, we provide a graphical explanation to illustrate the implementation of the proposed algorithm.

#### REMARK 1

We implemented the proposed algorithm with R calling C programming language, which is a few dozen times faster than using R programming language only. Such implementation is necessary for the application of our algorithm to real-world settings where the number of patients in each site is relatively large.

#### REMARK 2

In the situation that each site is treated as the local site, each site can obtain its own surrogate pairwise likelihood estimate. These estimates can be further synthesized together with the inverse variance weighted average method to obtain an overall estimate.

### Simulation Design

To evaluate the empirical performance of the proposed algorithm, we conduct a simulation study to cover a wide spectrum of practical settings. We set the total number of sites, K = 5 or 20 and sample size of each site is 1000 (i.e., the total numbers of patients are 5000 and 20,000 respectively.)

In our simulation study, we consider a setting where a binary outcome is associated with two risk factors, (*x*_1_,*x*_2_), where *x*_1_ represents a continuous predictor (e.g., age) and *x*_2_ is a binary predictor (e.g., sex, race). The binary outcome Y (e.g., presence/absence of hospitalization) is generated from a Bernoulli distribution, with the conditional probability specified by the following logistic regression model,

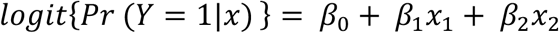

where *β*_1_ and *β*_2_ are the coefficients of *x*_1_ and *x*_2_ respectively, and *β*_0_ is the intercept, characterizing the prevalence of the outcome Y. We set the true value of *β*_1_ is 1 and of *β*_2_ is -1. The distribution of *x*_1_ for each study site is *Uni* (−1,1) to mimic the empirical distribution of variable “age”, and *x*_2_ is generated from a Bernoulli distribution with probability equal to 0.5 to mimic the empirical distribution of variable “sex”.

We simulated three scenarios of the disease prevalence. The medians of the prevalence are 20%, 5%, and 0.5%. Specifically, the prevalence of the sites is randomly generated from a range of values as presented in **Figure 3**. We also simulated two scenarios of heterogeneity under each disease prevalence to mimic less heterogeneous cases (upper panel) and more heterogeneous (lower panel) cases, where the prevalence ranges are larger than those of the less heterogeneous cases.

**Figure 2:**
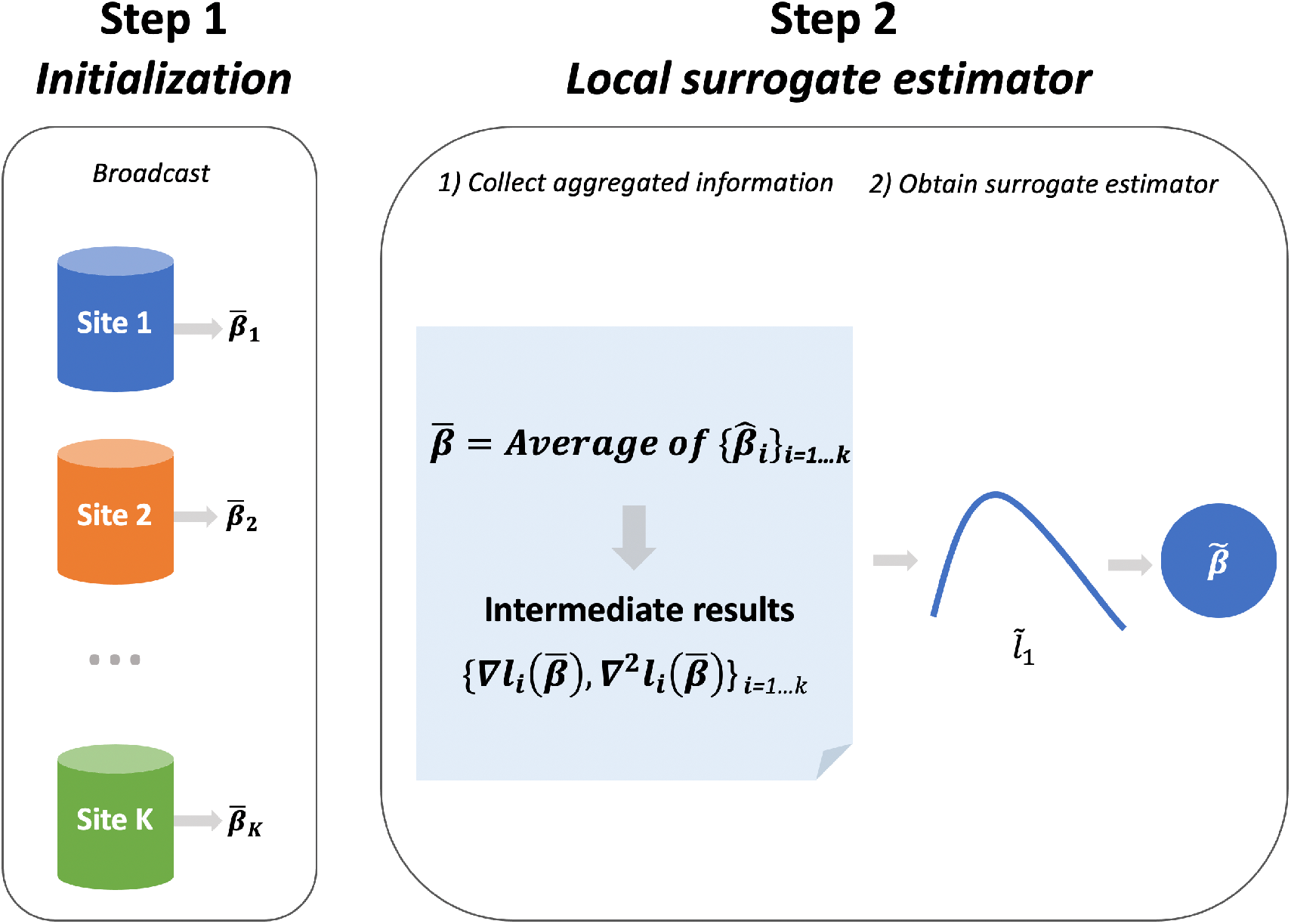
Illustration of the proposed method. Step I: Using data from each local site to estimate 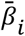, where *i* = 1,…,*K* and broadcast the values to calculate the weighted initial value 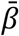. Step II: With 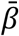, calculating the intermediate terms 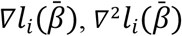 at each site and then transfer the results back to the local site. With the intermediate results and 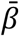 to construct the surrogate pairwise log-likelihood function 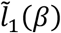 in the local site. Maximizing 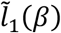 to obtain the estimator 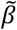.

**Figure 3:**
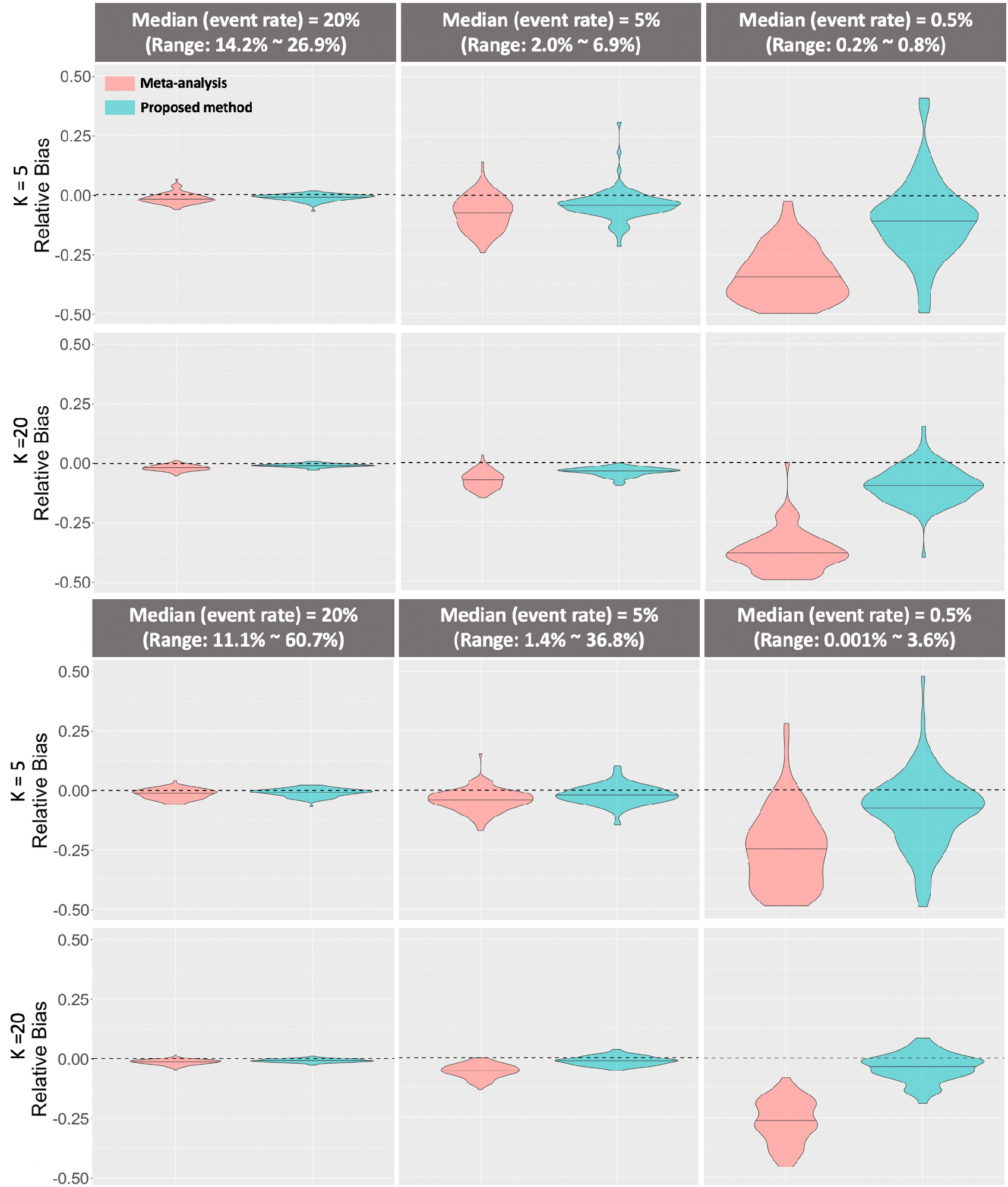
**Upper panel**: relative bias of *β*_1_ estimation compared with the pairwise likelihood method under three scenarios with median prevalence 20%, 5%, and 0.5% when the total number of sites is 5 or 20 (i.e., K = 5 or 20). **Lower panel**: relative bias of *β*_1_ estimation compared with the pairwise likelihood method under three scenarios with median prevalence 20%, 5%, and 0.5% with larger heterogeneity (i.e., larger prevalence range than upper panel) when the total number of sites is 5 or 20 (i.e., K = 5 or 20).

Under each scenario, we compared the proposed method with the pairwise likelihood method (Liang, 1987), which can be treated as the gold standard and the commonly used meta-analysis. In the pairwise likelihood method, we assume that we have the access to all of the patient-level data. The simulation was conducted with 100 replications.

### Data Evaluation

Our analytical dataset is composed of hospitalized patients who were diagnosed with COVID-19 prior to June 29, 2020 from a single large national health insurer, which covers a broad swath of the population. The data are from multiple EHR systems including EPIC [34], Cerner [35], and others. The data are recorded from *K* = 230 sites with 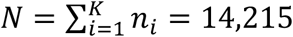 insured (Commercial and Medicare) patients; see **Figure 4 (a)** for the details of inclusion-exclusion criteria and the distribution of COVID patients across the United States. Our objective is to develop an association model between clinical-and-demographic covariates (i.e., age, sex, line of business, and Charlson comorbidity index) and therapeutic patient outcomes. More details about the data quality are provided in Supplementary Appendix 2.

**Figure 4:**
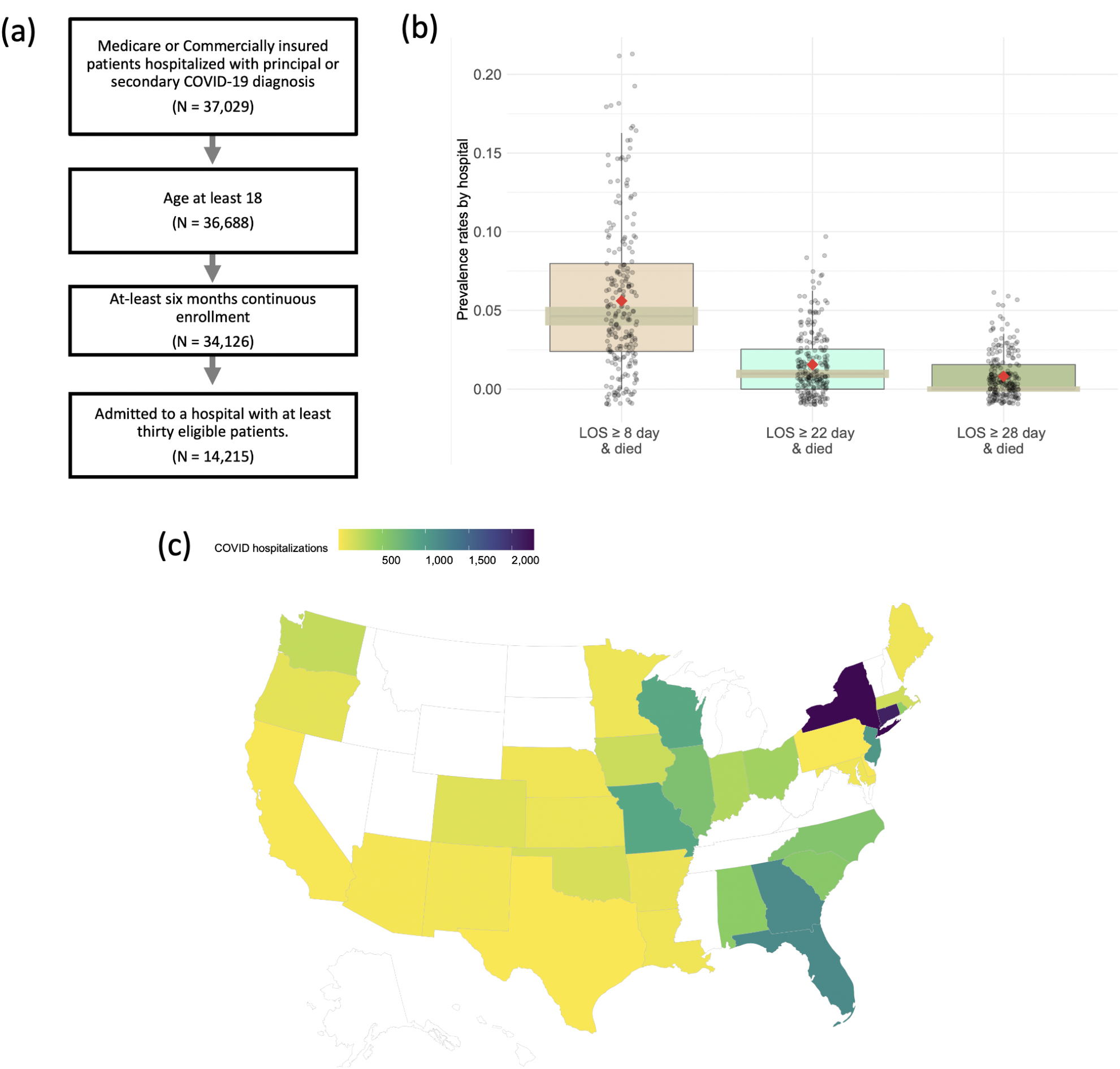
(a) Diagram of the patient inclusion-exclusion criteria. (b) Box plots of the prevalence rates of composite outcomes of 230 hospitals. (c) COVID-19 cases distribution: number of COVID-19 hospitalizations included in the study are represented across 47 states.

Outcomes are defined by combining both hospitalizations (days) and the status of patients being expired (i.e., a binary value taking value 1 if a patient is deceased or 0 otherwise). Consider three composite binary outcomes which take values 1 if the event occurs, and 0, otherwise. Here the events are defined as (a) LOS > 1 week and patient died, (b) LOS > 3 weeks and patient died, and (c) LOS > 4 weeks and patient died, respectively. **Figure 4 (b)** illustrates the prevalence rates of composite outcomes by 230 hospitals and **Figure 4 (c)** shows the number of COVID-19 hospitalizations included in the study across 47 states in the U.S. These two figures exhibit substantial variation in prevalence rates across sites. Moreover, patients admitted within the same hospital are subject to somewhat similar care, administrative facilities, and treatments provided by the same physicians. This phenomenon leads us to treat sites as internally homogeneous and externally heterogeneous blocks. For details of the covariates, we refer to **Table 1**.

**Table 1.** Summary characteristics of the 14,215 patients from 230 hospitals in our population.

|  |  |  |
| --- | --- | --- |
| **Number of patients** |  | 14,215 |
| **Number of hospitals** |  | 230 |
| **Patient Level Characteristics** |  |  |
|  | **Mean Age in years (Median, SD)** | 71.1 (73, 14.3) |
|  | **Sex** |  |
|  | Male (%) | 6,925 (48.7%) |
|  | Female (%) | 7,290 (51.3%) |
|  | **Mean Charlson Score (Median, SD)** | 3.4 (3.0, 3.0) |
|  | **Insurance Type** |  |
|  | Medicare Advantage (%) | 11,460 (80.6%) |
|  | Commercial (%) | 2,755 (19.4%) |
| **Patient Outcomes** |  |  |
|  | **Mean Length of Stay in days (Median, SD)** | 10.2 (6, 12.6) |
| | Length of Stay ≥ 1 day and Died (%) | 1,716 (12.7%) |
| | Length of Stay ≥ 8 day and Died (%) | 843 (5.9%) |
| | Length of Stay ≥ 15 day and Died (%) | 436 (3.1%) |
| | Length of Stay ≥ 22 day and Died (%) | 234 (1.6%) |
| | Length of Stay ≥ 29 day and Died (%) | 124 (0.9%) |

## RESULTS

### Simulation studies results

For simplicity, we only present the results for the estimation of coefficient *β*_1_ and the results for the other coefficients are similar. **Figure 3** shows the violin plot of the relative bias compared with the pairwise likelihood method under different numbers of sites and event rates. The first row in each panel is for the results when the total number sites, K = 5, and the second row is for K = 20. The black dashed line represents zero relative bias compared with the gold standard method. From the figure we observe that for all scenarios, the proposed method obtains smaller relative bias compared with meta-analysis. Importantly, as the event rate decreases under both less and more heterogeneous cases, the meta-analysis estimator is observed to have larger bias. When the event rate is <5%, the relative bias of the proposed estimator is 30% smaller than that of the meta-analysis estimator. In summary, the proposed method can provide better performance than the meta-analysis estimators to handle the heterogeneity across the clinical sites when the event is rare.

### Data evaluation results

We primarily focus on estimating and comparing parameter estimates by the proposed method and the meta-analysis method. We stress that the parameter estimates need to be interpreted with caution since the effects’ magnitudes or directions might be misleading without adjusting for potential confounders in the model. **Figure 5** illustrates the results obtained by the pairwise likelihood method (i.e., gold standard), the proposed method, and meta-analysis. As the prevalence rate decreases (i.e., in rare events), the proposed method outperforms meta-analysis in terms of estimating parameters. Specifically, the odds ratio (OR) of the proposed method remains closer to that of the gold standard approach, compared with the OR of meta-analysis. The proposed estimates have a relative bias <9% when the event rate is <1%, whereas the meta-analysis estimates have a relative bias at least 10% higher than that of the proposed method. This observation matches with that of the simulation study.

**Figure 5:**
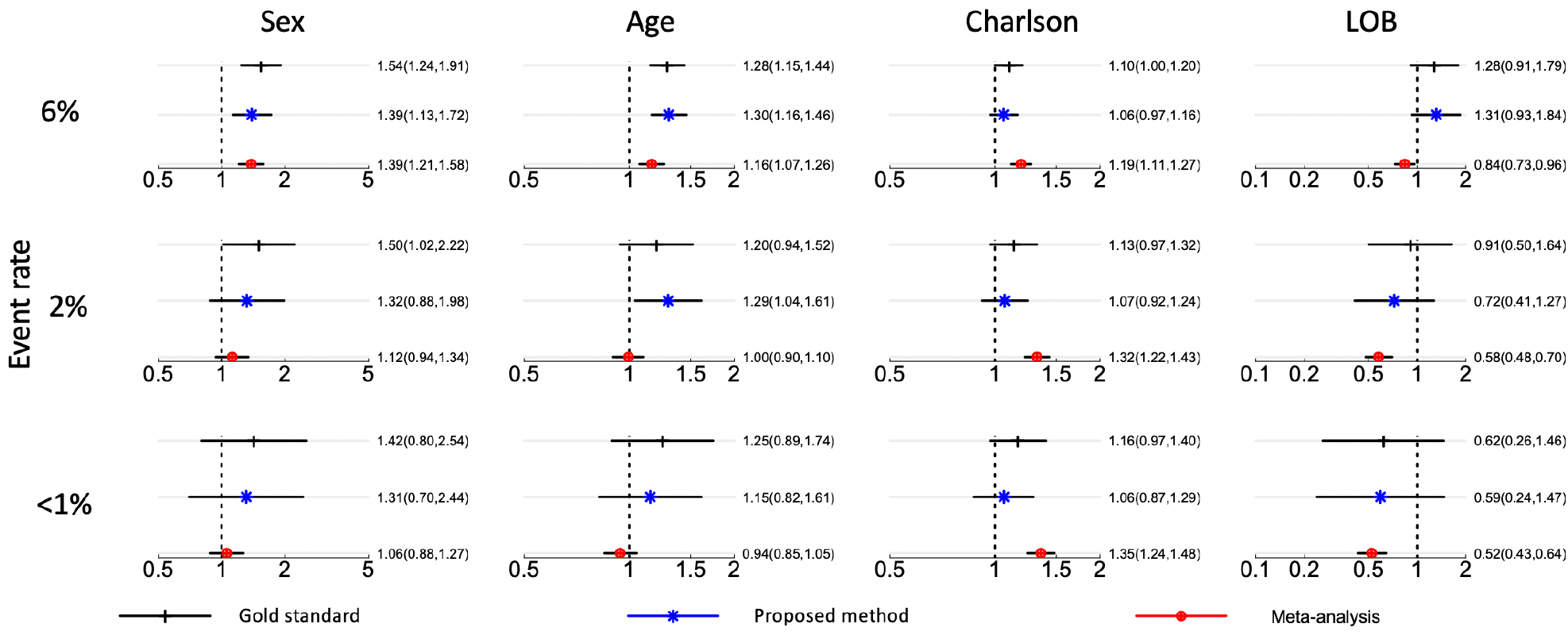
Point estimates and 95% confidence intervals (CI) for the association (in odds ratio scale) between the LOS (i.e., length of stay) and covariates (i.e., sex, age, Charlson score, line of business, from left to right). Each row represents an event rate of the outcome: 6%, 2%, and <1% from top to bottom. Each column represents the estimation of the covariate.

Besides, meta-analysis underestimates variance (or standard error of estimates) leading to far narrower confidence intervals relative to those of the gold standard method, especially for rare events. Ignoring between-and-within sites correlation in meta-analysis is likely to induce bias and underestimate uncertainty in parameter estimates leading to conflicting inference about the testing of significance of the effect size. For example, 95% confidence intervals of ORs for Charlson score based on meta-analysis does not contain OR value of one implying its significance, which is inconsistent with the inference based on the gold standard method. In contrast, the proposed method produces comparable inferences to the gold standard method.

## DISCUSSION

In this paper, we proposed an effective privacy-preserving distributed algorithm for modeling binary outcomes while accounting for heterogeneity across clinical sites. Motivated by real-world multicenter data, the proposed method requires transferring initial values and intermediate information instead of patient-level data. Our algorithm provides an estimator that is robust to heterogeneity in event rates. In simulations, the proposed method is shown to have higher accuracy than meta-analysis when the outcome is relatively rare, suggesting its utility in a rare-event context.

There are several advantages of our proposed algorithm compared to existing methods for privacy-preserving data analysis. Relative to meta-analysis, our method accesses patient data at a higher granularity while requiring minimal additional effort to institute. For multi-site studies operating under a common data model, such as OHDSI, analyses using our method can be carried out at individual sites concurrently without the need for any site-specific modifications. In addition, there are many benefits of using our method compared to existing distributed algorithms. First, compared to the iterative algorithms such as GLORE and WebDISCO [26,27], the proposed algorithm does not require iterative communication across the sites, leading to the reduction in communication costs and administrative efforts. Secondly, to implement the proposed method, the patient-level data are only required in one single site. For the other sites within the network, the aggregated information will be used instead of patient-level data transfer across the sites to construct the surrogate pairwise likelihood function. Given the understandable privacy- and proprietary-related sensitivities health systems have to provide “outside” collaborators with access to patient-level data, limiting the need to use such data to only one site would be extremely beneficial to a multi-site project in terms of feasibility, costs, and time. Thirdly, by canceling out the baseline probability function, the proposed method can handle the heterogeneity in the event rate between the sites. In addition, the proof of the proposed method is established based on U statistics and is different from that of Jordan et al. (2019) [33]. In terms of computational complexity, the implementation of the proposed method is slow when the sample size is large compared with the traditional regression model. We thus implemented the algorithm with R calling C, which is a few dozen times faster than using the R programming language only. The R and C code will be made available at https://github.com/Penncil and through our R package: ‘pda’.

To investigate the proposed non-iterative distributed algorithm, we can extend it in several aspects. First, the proposed pairwise likelihood function can only handle the heterogeneity of the intercepts in the regression model. To handle the other types of heterogeneity (e.g., heterogeneous effects of the predictors), more robust algorithms should be developed. Secondly, we are going to develop methods for other types of outcomes, such as continuous and time-to-event data. In addition, the development of distributed algorithms to handle the missingness in the longitudinal EHR is needed in the future. Lastly, we have been working on the development of the open-source software R package to implement the proposed distributed algorithm within a multicenter network. We believe that the proposed algorithm would be a robust method to account for the heterogeneity across multiple clinical sites, leading to a better data integration framework inside health systems.

## CONCLUSION

The proposed distributed algorithm provides an estimator that is robust to heterogeneity in event rates when effectively integrating data from multiple clinical sites. Through a simulation study and a real-world use case using data from the UnitedHealth Group Clinical Research Database, the proposed method is shown to be an effective alternative to both meta-analysis and existing distributed algorithms for modeling heterogeneous multi-site binary outcomes.

## Data Availability

The dataset is composed of 14,215 hospitalized patients who were diagnosed with COVID-19 prior to June 29, 2020 from a single large national health insurer at UnitedHealth Group Clinical Research Database, which covers a broad swath of the population. The data are from multiple EHR systems including EPIC, Cerner, and others. The data are proprietary and are not available for public use but can be made available to editors and their approved auditors under a data use agreement to confirm the findings of the current study.

## AUTHOR CONTRIBUTIONS

JT and YC designed methods and experiments; MI, NS, and JN provided the dataset from UnitedHealth Group Clinical Research Database for data analysis; CL, RD and YC guided the theoretical development and dataset generation for the simulation study; JT generated the simulation datasets, conducted simulation experiments; MI, NS and JB conducted data analysis of the EHR data from the UnitedHealth Group; all authors interpreted the results and provided instructive comments; JT drafted the main manuscript. All authors have approved the manuscript.

