## Supplementary Appendix for "An efficient distributed algorithm with application to COVID-19 data from heterogeneous clinical sites"

October 16, 2020

### Summary

In this supplemental file, we provide the following materials:

1): Statistical Inference and Proof

2): Database Quality

[1]Perelman School of Medicine, The University of Pennsylvania, Philadelphia, PA, USA
[2]UnitedHealth Group, Minnetonka, MN, USA
[3]School of Public Health, Harvard University, MA, USA
*Corresponding authors: Yong Chen, University of Pennsylvania, Perelman School of Medicine, Blockley Hall 602, 423 Guardian Drive, Philadelphia, PA 19104, Office: 215-746-8155,

### Appendix 1. Statistical Inference and Proof

#### *1.1 Theorem*

Let $\hat{\beta}$ denote the maximum pairwise likelihood function estimator of $L^*(\beta)$ defined in equation (3). The following proposition gives the asymptotic distribution of the pairwise likelihood estimator $\hat{\beta}$, which has been established in Liang (1987).

PROPOSITION 1: With $K$ fixed and $n$ increased, we have

$$\sqrt{Kn}(\hat{\beta} - \beta) \rightarrow N(0, V) \tag{A.1}$$

where $V = {\Sigma_1}^{-1}\Sigma_2{\Sigma_1}^{-1}$, and

$$\Sigma_1 = E(\frac{\partial S^{(jl)}(\beta)}{\partial\beta}), \Sigma_2 = cov\{S^{(jl)}(\beta)\} \tag{A.2}$$

where $S^{(jl)}(\beta) = \sum_{i=1}^{K} {S_i}^{(jl)}(\beta)/K$ and ${S_i}^{(jl)}(\beta) = (y_{ij} - y_{il})(x_{ij} - x_{il})exp\{-(y_{ij} - y_{il})(x_{ij} - x_{il})^T\beta\}/(1 + exp\{-(y_{ij} - y_{il})(x_{ij} - x_{il})^T\beta\})$. Build on Liang (1987), we now provide the large sample distribution of the proposed surrogate estimator.

THEOREM 1: *As $n$ increases, given the initial estimator $\bar{\beta}$ satisfies $||\bar{\beta} - \hat{\beta}|| = O(n^{-1/2})$, the proposed surrogate pairwise likelihood estimator $\tilde{\beta}$ satisfies*

$$\sqrt{n}||\tilde{\beta} - \hat{\beta}|| = O(n^{-1/2})\ ||\bar{\beta} - \hat{\beta}||, \tag{A.3}$$

*where $\hat{\beta}$ is the maximum pairwise likelihood estimator of $L^*(\beta)$.*

Since the pairs $\{(y_{ij,}y_{il}), 1 \leq j < l \leq n\}$ consisting the pairwise likelihood are not i.i.d, the proof of Theorem 1 is based on the U statistics and is different from that of Jordan et al. (2019)

REMARK 1: Theorem 1 implies that the surrogate estimate $\tilde{\beta}$ has the same limiting distribution as $\hat{\beta}$. In a distributed setting, the variance of $\tilde{\beta}$ can be consistently estimated by its empirical estimator $\hat{\Sigma}_{1,i=1}{}^{-1}(\tilde{\beta})\hat{\Sigma}_{1,i=1}(\tilde{\beta})\hat{\Sigma}_{1,i=1}{}^{-1}(\tilde{\beta})/\binom{n}{2}$ with the patient-level data from the local site (i.e., site 1; i = 1), where and are function of $\beta$ for the i-th site,

$$\hat{\Sigma}_{1,i}(\beta) = \binom{n}{2}^{-1} \sum_{j<l} - \frac{\partial s_i{}^{(jl)}(\beta)}{\partial \beta}, \tag{A.4}$$

$$\hat{\Sigma}_{2,i}(\beta) = \frac{4}{n(n-1)(n-2)} \sum_{j<l} S_i{}^{(jl)}(\beta) S_i{}^{(jl)}(\beta)^T, \tag{A.5}$$

There is no extra communication across the sites after we obtain the $\tilde{\beta}$ in the calculation of variance of $\tilde{\beta}$.

#### *1.2 Proof of Theorem 1*

In the proof below, we rewrite the log-conditional likelihood term

$$\ell_i^{jl} = \log\left\{1 + \exp\left(\left(y_{ij} - y_{il}\right)\left(x_{ij} - x_{il}\right)\beta^T\right)\right\}.$$

We denote $\|.\|$ the L-2 norm for a vector or Frobenius norm for a matrix. Before proving Theorem 1, we first introduce the regularity conditions and some corollaries. First of all, we present a result about the Lipschitz continuity of $\nabla^2 \ell_i^{jl}$.

COROLLARY 1: *For any $i, j, \beta, \beta'$, we have*

$$\|\nabla^2 \ell_i^{jl}(\beta') - \nabla^2 \ell_i^{jl}(\beta)\| \leq L(X_{ij}, Y_{ij}, X_{il}, Y_{il}) \|\beta' - \beta\|$$

*where $L(X_{ij}, Y_{ij}, X_{il}, Y_{il})$ is given by*

$$L\left(X_{ij}, Y_{ij}, X_{il}, Y_{il}\right) = \left\|Y_{ij} - Y_{il}\right\|^3 \left\|X_{ij} - X_{il}\right\|^3.$$

*Proof:* Based on the likelihood, we have

$$\nabla^2 \ell_i^{jl}(\beta') - \nabla^2 \ell_i^{jl}(\beta) = \left\{\frac{T'}{(1+T')^2} - \frac{T}{(1+T)^2}\right\} \left(y_{ij} - y_{il}\right)^2 \left(x_{ij} - x_{il}\right)^{\otimes 2}$$

where $T' = \exp\left(\left(y_{ij} - y_{il}\right)\left(x_{ij} - x_{il}\right)'\beta'\right)$ and $T = \exp\left(\left(y_{ij} - y_{il}\right)\left(x_{ij} - x_{il}\right)'\beta\right)$. So,

$$
\begin{aligned}
&\left\|\nabla^2 \ell_i^{jl}(\beta') - \nabla^2 \ell_i^{jl}(\beta)\right\| \\
&\leq \left|\frac{T'}{(1+T')^2} - \frac{T}{(1+T)^2}\right| \left\|\left(y_{ij} - y_{il}\right)^2 \left(x_{ij} - x_{il}\right)^{\otimes 2}\right\| \\
&= \left\|\left(y_{ij} - y_{il}\right)^2 \left(x_{ij} - x_{il}\right)^{\otimes 2}\right\| \left|\frac{(T'-T)(1-T'T)}{(1+T')^2(1+T)^2}\right| \\
&\leq \left\|\left(y_{ij} - y_{il}\right)^2 \left(x_{ij} - x_{il}\right)^{\otimes 2}\right\| \left|\frac{(T'-T)}{(1+T')(1+T)}\right| \left|\frac{(1-T'T)}{(1+T')(1+T)}\right| \\
&\leq \left\|\left(y_{ij} - y_{il}\right)^2 \left(x_{ij} - x_{il}\right)^{\otimes 2}\right\| \left|\frac{(T'-T)}{(1+T')(1+T)}\right| \\
&\leq \left|y_{ij} - y_{il}\right|^3 \|x_{ij} - x_{il}\|^3 \left|\frac{T''}{(1+T')(1+T)}\right| \|\beta' - \bar{\beta}\|
\end{aligned}
$$

with

$$
T'' = \exp\left(\left(y_{ij} - y_{il}\right)\left(x_{ij} - x_{il}\right)'\beta''\right) = (T')^{a'}(T)^{1-a'} \leq \max\{T', T\},
$$

and $\beta'' = a'\beta' + (1-a')\beta$ is the linear combination of $\beta$ and $\beta'$. $\beta''$ is between $\beta$ and $\beta'$, $0 \leq a' \leq 1$. Thus,

$$
\left\|\boldsymbol{\nabla^2 \ell_i^{jl}(\beta') - \nabla^2 \ell_i^{jl}(\beta)}\right\| \leq \left|\boldsymbol{y_{ij} - y_{il}}\right|^3 \|\boldsymbol{x_{ij} - x_{il}}\|^3 \|\boldsymbol{\beta' - \bar{\beta}}\|
$$

Then, we introduce the following conditions for Theorem 1.

Condition 1 (**Local convexity**): The Hessian matrix $I_i(\beta) = \mathbb{E}_i[\nabla^2 \ell_j(\beta)]$ of the pairwise log-likelihood function $\ell_i$ at site I is invertible at $\hat{\beta}$, there exist positive constant $\lambda$, such as $\lambda I_p \preccurlyeq \nabla^2 \ell_j(\beta)$.

Condition 2 (**Smoothness**): There exist constants $(G, H)$ such that $\mathbb{E}\left\|\nabla\ell_j^{1,2}(\beta)\right\|_2^4 \leq G^4$,

$\mathbb{E}\left\|\nabla\ell_j^{1,2}(\beta) - I_i(\beta)\right\|_2^4 \leq H^4$ for all $k$ and $\beta \in U(\rho) = \{\beta: \|\beta - \hat{\beta}\} \leq \rho$. For the function $L(X_1, Y_1, X_2, Y_2)$ in Corollary 1, there exist constant L such that $\mathbb{E}_i[L(X_1, Y_1, X_2, Y_2)^2] < L^2$ and $\mathbb{E}_i[\{L(X_1, Y_1, X_2, Y_2) - \mathbb{E}_i[L(X_1, Y_1, X_2, Y_2)]\}^2] < L^2$.

COROLLARY 2: *The following inequality holds with probability* $1 - O(n^{-1})$.

$$\frac{1}{K\binom{n}{2}}\sum_{i,j,l} L(X_{ij}, Y_{ij}, X_{il}, Y_{il}) \leq 4\,\mathbb{E}L(X_{ij}, Y_{ij}, X_{il}, Y_{il}) + 6L$$

*Proof:* Define $\tilde{L}(X_{ij}, Y_{ij}, X_{il}, Y_{il}) = L(X_{ij}, Y_{ij}, X_{il}, Y_{il}) - \mathbb{E}L(X_{ij}, Y_{ij}, X_{il}, Y_{il})$. Now we calculate the following probability of the inequality using Markov inequality:

$$\mathbb{P}\left(\frac{1}{K\binom{n}{2}}\sum_{i,j,l} L(X_{ij}, Y_{ij}, X_{il}, Y_{il}) > 4\,\mathbb{E}L(X_{ij}, Y_{ij}, X_{il}, Y_{il}) + 6L\right)$$

$$= \mathbb{P}\left(\frac{1}{K\binom{n}{2}}\sum_{i,j,l} \tilde{L}(X_{ij}, Y_{ij}, X_{il}, Y_{il}) > 3\,\mathbb{E}L(X_{ij}, Y_{ij}, X_{il}, Y_{il}) + 6L\right)$$

$$\leq \{3\mathbb{E}L(X_{ij}, Y_{ij}, X_{il}, Y_{il}) + 6L\}^{-2} Var\left(\frac{1}{K\binom{n}{2}}\sum_{i,j,l} \tilde{L}(X_{ij}, Y_{ij}, X_{il}, Y_{il})\right)$$

$$\leq \{3\mathbb{E}L(X_{ij}, Y_{ij}, X_{il}, Y_{il}) + 6L\}^{-2} \frac{4}{K^2 n^2 (n-1)^2} \{\sum_{i,j,l} Var\left(\tilde{L}(X_{ij}, Y_{ij}, X_{il}, Y_{il})\right)$$

$$+ \sum_{i,j,l,l'} Cov\left(\tilde{L}(X_{ij}, Y_{ij}, X_{il}, Y_{il}), \tilde{L}(X_{ij}, Y_{ij}, X_{il'}, Y_{il'})\right)\}$$

Now, let's prove Theorem 1.

*Proof*. Define the "good event"

$$\mathcal{E}_0 = \{\frac{1}{\binom{n}{2}} \sum L(X_i, X_j, Y_i, Y_j) \le 2L\},$$

$$\mathcal{E}_1 = \{\|\nabla^2 \tilde{\ell}_1(\hat{\beta}) - \nabla^2 \ell(\beta)\| \le \frac{\rho\lambda}{2}\},$$

$$\mathcal{E}_2 = \{\|\nabla \tilde{\ell}_1(\hat{\beta})\| \le \frac{(1-\rho)\lambda\delta_p}{4}\}$$

By Lemma 6 in Zhang et al. (2013), we have under event $\mathcal{E} = \mathcal{E}_0 \cap \mathcal{E}_1 \cap \mathcal{E}_2$, $\|\bar{\beta} - \hat{\beta}\| \le \frac{2\|\nabla \tilde{\ell}_1(\hat{\beta})\|}{(1-\rho)\lambda}$.

We then prove that $\|\nabla \tilde{\ell}_1(\hat{\beta})\| = O\left(n^{-\frac{1}{2}}\right) \|\bar{\beta} - \hat{\beta}\|$.

$$\begin{aligned}
\nabla \tilde{\ell}_1(\hat{\beta}) &= \nabla \ell_1(\hat{\beta}) + \{\nabla \ell(\bar{\beta}) - \nabla \ell_1(\bar{\beta})\} + \{\nabla^2 \ell(\bar{\beta}) - \nabla^2 \ell_1(\bar{\beta})\}(\hat{\beta} - \bar{\beta}) - \nabla \ell(\hat{\beta}) \\
&= \{\nabla \ell_1(\hat{\beta}) - \nabla \ell_1(\bar{\beta})\} + \{\nabla \ell(\hat{\beta}) - \nabla \ell(\bar{\beta})\} + \{\nabla^2 \ell(\bar{\beta}) - \nabla^2 \ell_1(\bar{\beta})\}(\hat{\beta} - \bar{\beta}) \\
&= -\nabla^2 \ell_1(\beta')(\bar{\beta} - \hat{\beta}) + \nabla^2 \ell(\beta')(\bar{\beta} - \hat{\beta}) - \{\nabla^2 \ell(\bar{\beta}) - \nabla^2 \ell_1(\bar{\beta})\}(\bar{\beta} - \hat{\beta}) \\
&= \{\nabla^2 \ell(\beta') - \nabla^2 \ell_1(\beta') - \nabla^2 \ell(\bar{\beta}) + \nabla^2 \ell_1(\bar{\beta})\}(\bar{\beta} - \hat{\beta})
\end{aligned}$$

where $\beta' = a\,\hat{\beta} + (1-a)\ \bar{\beta}, 0 \le a \le 1$. The first equation is due to the definition of $\tilde{\ell}_1$ and $\nabla \ell(\hat{\beta}) = 0$. Therefore,

$$\|\nabla \tilde{\ell}_1(\hat{\beta})\| \le \{\|\nabla^2 \ell(\beta') - \nabla^2 \ell(\bar{\beta})\| + \|\nabla^2 \ell_1(\beta') - \nabla^2 \ell_1(\bar{\beta})\|\}\|\bar{\beta} - \hat{\beta}\|$$

Based on the smoothness Condition 2 and Corollary 2, we have

$$\begin{aligned}
\|\nabla \tilde{\ell}_1(\hat{\beta})\| \le &\left\{\left\|\sum_{i,j,l} \frac{1}{K\binom{n}{2}} \left[\nabla^2 \ell_i^{jl}(\beta') - \nabla^2 \ell_i^{jl}(\bar{\beta})\right]\right\|\right. \\
&\left. + \left\|\sum_{j,l} \frac{1}{\binom{n}{2}} \left[\nabla^2 \ell_1^{jl}(\beta') - \nabla^2 \ell_1^{jl}(\bar{\beta})\right]\right\|\right\} \|\bar{\beta} - \hat{\beta}\| \\
\le &\left[\frac{1}{K\binom{n}{2}} \sum_{i,j,l} L(X_{ij}, Y_{ij}, X_{il}, Y_{il}) + \frac{1}{\binom{n}{2}} \sum_{j,l} L(X_{ij}, Y_{ij}, X_{il}, Y_{il})\right] \|\beta' - \bar{\beta}\| \|\bar{\beta} - \hat{\beta}\|
\end{aligned}$$

$$\leq \left(4\mathbb{E}L(X_{ij}, Y_{ij}, X_{il}, Y_{il}) + 6L\right)\|\beta' - \bar{\beta}\|\|\bar{\beta} - \hat{\beta}\|$$

$$= O(\mathrm{n}^{-1/2})\|\bar{\beta} - \hat{\beta}\|$$

We get the last equation because $\|\beta' - \bar{\beta}\| \leq \|\bar{\beta} - \hat{\beta}\| = O(n^{-1/2})$. Then, we need to prove the high probability bound of the "good events". First, we investigate the event $\mathcal{E}_0$. The probability of the $\mathcal{E}_0^c$ can be calculated with Markov inequality as follows:

$$\mathbb{P}(\mathcal{E}_0^c) \leq \mathbb{P}\left(\frac{1}{\binom{n}{2}}\sum_{j,l} \tilde{L}(X_{ij}, Y_{ij}, X_{il}, Y_{il}) > -\mathbb{E}L(X_{ij}, Y_{ij}, X_{il}, Y_{il}) + 2L\right)$$

$$\leq \left[-\mathbb{E}L(X_{ij}, Y_{ij}, X_{il}, Y_{il}) + 2L\right]^{-2} Var\left(\frac{1}{\binom{n}{2}}\sum_{j,l} \tilde{L}(X_{ij}, Y_{ij}, X_{il}, Y_{il})\right)$$

$$\leq \left[-\mathbb{E}L(X_{ij}, Y_{ij}, X_{il}, Y_{il}) + 2L\right]^{-2} \frac{1}{n^2(n-1)^2}\left\{\sum_{j,l} Var\tilde{L}(X_{ij}, Y_{ij}, X_{il}, Y_{il})\right.$$

$$\left. + \sum_{j,l,l'} Cov\left(\tilde{L}(X_{ij}, Y_{ij}, X_{il}, Y_{il}), \tilde{L}(X_{ij}, Y_{ij}, X_{il'}, Y_{il'})\right)\right\}$$

$$\leq O(n^{-1})$$

We then investigate $\mathcal{E}_1$. Then, we have

$$\mathbb{P}(\mathcal{E}_1^c) = \mathbb{P}\left(\|\nabla^2\ell_1(\hat{\beta}) + \nabla^2\ell(\bar{\beta}) - \nabla^2\ell_1(\bar{\beta}) - \nabla^2\ell(\hat{\beta})\| > \frac{\rho\lambda}{2}\right)$$

$$\leq \mathbb{P}\left(\frac{1}{\binom{n}{2}}\|\sum_{j,l} \nabla^2\ell_1^{jl}(\hat{\beta}) - \nabla^2\ell_1^{jl}(\bar{\beta})\| > \frac{\rho\lambda}{4}\right) + \mathbb{P}\left(\frac{1}{K\binom{n}{2}}\|\sum_{i,j,l} \nabla^2\ell_k^{jl}(\hat{\beta}) - \nabla^2\ell_k^{jl}(\bar{\beta})\| > \frac{\rho\lambda}{4}\right)$$

Let $\beta' = \hat{\beta}$ and $\beta = \bar{\beta}$ in Corollary 1, then we have

$$\|\nabla^2\ell_i^{jl}(\hat{\beta}) - \nabla^2\ell_i^{jl}(\bar{\beta})\| \leq L(X_{ij}, Y_{ij}, X_{il}, Y_{il})\|\hat{\beta} - \bar{\beta}\|$$

In the above function, the first term has the following probability for large enough n,

$$\mathbb{P}\left(\frac{1}{\binom{n}{2}}\left\|\sum_{j,l}\nabla^2\ell_1^{jl}(\hat{\beta})-\nabla^2\ell_1^{jl}(\bar{\beta})\right\|>\frac{\rho\lambda}{4}\right)$$

$$\leq\mathbb{P}\left(\frac{1}{\binom{n}{2}}\sum_{j,l}\left\|\nabla^2\ell_1^{jl}(\hat{\beta})-\nabla^2\ell_1^{jl}(\bar{\beta})\right\|>\frac{\rho\lambda}{4}\right)$$

$$\leq\mathbb{P}\left(\frac{1}{\binom{n}{2}}\sum_{j,l}L\left(X_{1j},Y_{1j},X_{1l},Y_{1l}\right)>\frac{\rho\lambda}{4\|\hat{\beta}-\bar{\beta}\|}\right)$$

$$\leq\mathbb{P}\left(\frac{1}{\binom{n}{2}}\sum_{j,l}L\left(X_{1j},Y_{1j},X_{1l},Y_{1l}\right)>2L\right)$$

$$=\ \mathbb{P}(\mathcal{E}_0^c)=O(n^{-1})$$

Similarly, for the second term, we have:

$$\mathbb{P}\left(\frac{1}{K\binom{n}{2}}\left\|\sum_{i,j,l}\nabla^2\ell_i^{jl}(\hat{\beta})-\nabla^2\ell_i^{jl}(\bar{\beta})\right\|>\frac{\rho\lambda}{4}\right)=O(n^{-1})$$

Thus, $\mathbb{P}(\mathcal{E}_1^c)=O(n^{-1})$. Finally, we can investigate $\mathcal{E}_2$.

$$\mathbb{P}(\mathcal{E}_2^c)=\mathbb{P}\left(\left\|\nabla\,\tilde{\ell}_1(\hat{\beta})\right\|>\frac{(1-\rho)\lambda\delta_p}{4}\right)$$

$$\leq\mathbb{P}\left(\left\{\left\|\nabla^2\ell(\beta')-\nabla^2\ell(\bar{\beta})\right\|+\left\|\nabla^2\ell_1(\beta')-\nabla^2\ell_1(\bar{\beta})\right\|\right\}\left\|\beta'-\hat{\beta}\right\|>\frac{(1-\rho)\lambda\delta_p}{4}\right)$$

$$\leq\ \mathbb{P}\left(\left\{\left\|\nabla^2\ell(\beta')-\nabla^2\ell(\bar{\beta})\right\|\right\}>\frac{(1-\rho)\lambda\delta_p}{4\|\beta'-\hat{\beta}\|}\right)+\mathbb{P}\left(\left\{\left\|\nabla^2\ell_1(\beta')-\nabla^2\ell_1(\bar{\beta})\right\|\right\}>\frac{(1-\rho)\lambda\delta_p}{4\|\beta'-\hat{\beta}\|}\right)$$

$$\leq O(n^{-1})$$

**Appendix 2. Database Quality**

**Standardization of Data Entry and Data Structure**

Medical and pharmacy claims data are captured, predominantly electronically, from sites of care seeking third-party reimbursement for both Medicare and commercial plans using the industry standard data collection forms HCFA/CMS-1500 for facility claims, UB04/CMS-1450 for professional services and outpatient claims, and NCPDP for pharmacy claims or their electronic equivalents. Structured data from these standardized forms are coded using the International Classification of Diseases, Tenth Revision, Clinical Modification (ICD-10-CM), National Drug Codes (NDC), Current Procedural Terminology (CPT) codes, and Logical Observation Identifiers Names and Codes (LOINC) codes, and Diagnosis Related Groups (DRG). This nomenclature ensures consistency of data collection across geographic regions, health systems, and payers throughout the United States.

**Methods to Control for Errors in Sampling and Data Collection**

Claims that do not adhere to the form or coding standards described above are rejected from reimbursement, minimizing the risk that inappropriately structured data are included in the database. Data specific to SARS-CoV-2 and COVID-19 has an additional Quality Control layer to control for errors in sampling and data collection; this is described below in the section on Quality Control.

### Data Relevance and Accuracy

Data are transferred into the UnitedHealth Group (UHG) Clinical Discovery Database, where a dedicated team pursues data management to ensure accurate matching of source data to an individual. This protocol uses unique identifiers to match them to existing identifiers in the UHG Clinical Discovery Database to determine whether the individual already exists in the platform. A unique identification number is generated for each individual so that data from multiple sources can be linked back to that identification number. Individuals that fail to meet the matching criteria are excluded from the UHG Clinical Discovery Database to reduce the risk of erroneous linkage of records. Those whose claims do not fulfill basic standardized data structure requirements described previously are also excluded. During this, all member protected data are stored in a separate database that is only accessible by a designated engineering team. In addition to a persistent identifier being generated for each member, a de-identified primary key is also generated. The de-identified primary key is recycled every 6 months, at which time each member is assigned a new de-identified primary key. Data that are made available for research through the UHG Clinical Discovery Database use the de-identified primary key as the link across data tables. All protected information has been removed, ensuring any research performed is limited to retrospective analysis of de-identified data and accessed in accordance with Health Insurance Portability and Accountability Act regulations.

### Sufficiency of Basic Data

As described above, individuals lacking enough data to be assigned a unique primary key are excluded from the UHG Clinical Discovery Database, as are patients whose claims did not fulfill basic data structure requirements. In a given month in 2019, the UHG Clinical Discovery Database contained one or more claims from 5 million Medicare Advantage enrollees and 20 million commercially insured individuals. Further information on data sufficiency for the research performed in this manuscript can be found in **Figure 3 (a)** in the manuscript.

### Adequacy of Possible Derived Data

To reduce the risk of introducing error to standardized, structured claims data, derivation of source data within the UHG Clinical Discovery Database is minimal. The Data Integration team loads, formats, and join the data to appropriate dimension tables. Dimension tables are combined with raw claims information to limit the number of times external tables need to be referenced. Researchers may request derived fields within data tables prepared specifically for a project. This process is managed by the Data Enrichment team, who creates data dictionaries to accompany derived fields. Tables containing derived data are stored separately from raw source data.

### Design of Computer Editing Methods

Access to modify/edit source data is restricted to a subset of data specialists. Each step in the data flow has a restricted list of individuals able to perform any type of editing to the database, and access level varies by team (Data Integration, Data Enrichment). Researchers using the Clinical Discovery Database may not edit any source data or enrichment data. They are instead given access to “sandbox” locations where they may request editing access for the data tables used in their analyses.

**Quality Control**

In addition to the quality control mechanisms described during the matching procedures to reject non-linkable or inappropriately structured data, a COVID-19 data source-specific layer of quality control is also present, given the rapidly evolving situation. SARS-CoV-2 lab tests included in the UHG Clinical Discovery Database exclude custom local codes or codes that are not present in the LOINC organization's guidance for mapping SARS-CoV-2 and COVID-19 related LOINC terms. Test information provided via the LOINC code compliments the test type (antibody, RT-PCR, etc.) as well as the result value (detected, not detected, not given/cancelled). Suspected COVID-19 inpatient cases are manually reviewed daily by health plan clinical staff via clinical notes to determine an individual's COVID-19 status. Each case is then manually flagged as either negative, confirmed, presumed positive, or needs clinical review. If a case is confirmed, it is not reviewed again. If a case is listed as negative or unknown, it is periodically reviewed for changes in the record. All others are reviewed and updated daily.

**Differences Across Groups**

While the data for Medicare Advantage and commercially insured enrollees is processed in a similar manner, these groups are substantially different. First, there are systematic differences in patient characteristics, most remarkably the older age and the higher prevalence of all comorbidities. These differences are tabulated in the UHG Clinical Discovery Database, there are restrictions from individual employers on these use of data for research. Therefore, commercial insurance claims that are available for analyses are a subset of the overall commercially insured population.

**Data Sharing**

The data are proprietary and are not available for public use but can be made available to editors and their approved auditors under a data use agreement to confirm the findings of the current study.
